# Point-of-care sample preparation and automated quantitative detection of *Schistosoma haematobium* using mobile phone microscopy

**DOI:** 10.1101/2021.11.03.21265895

**Authors:** Maxim Armstrong, Andrew R. Harris, Michael V. D’Ambrosio, Jean T. Coulibaly, Samuel Essien-Baidoo, Richard K.D. Ephraim, Jason R. Andrews, Isaac I. Bogoch, Daniel A. Fletcher

## Abstract

*Schistosoma haematobium* continues to pose a significant public health burden despite ongoing global control efforts. One of several barriers to sustained control (and ultimately elimination) is the lack of access to highly sensitive diagnostic or screening tools that are inexpensive, rapid, and can be utilized at the point of sample collection. Here, we report an automated point-of-care diagnostic based on mobile phone microscopy that rapidly images and identifies *S. haematobium* eggs in urine samples. Parasite eggs are filtered from urine within a specialized, inexpensive cartridge that is then automatically imaged by the mobile phone microscope (the “SchistoScope”). Parasite eggs are captured at a constriction point in the tapered cartridge for easy imaging, and the automated quantification of eggs is obtained upon analysis of the images by an algorithm. We demonstrate *S. haematobium* egg detection with greater than 90% sensitivity and specificity using this device compared to the field gold standard of conventional filtration and microscopy. With simple sample preparation and image analysis on a mobile phone, the SchistoScope combines the diagnostic performance of conventional microscopy with the analytic performance of an expert technician. This portable device has the potential to provide rapid and quantitative diagnosis of *S. haematobium* to advance ongoing control efforts.

## Introduction

At least 100 million individuals in Africa are infected with *Schistosoma haematobium*, resulting in hundreds of thousands of deaths each year.^1^ The disability-adjusted life years (DALYs) lost and the economic burden of schistosomiasis places it among the most devastating diseases on the continent.^2^ *S. haematobium* causes urogenital disease and is responsible for a large spectrum of chronic illness, including chronic pelvic pain, infertility, and bladder cancer. Parasite control efforts consist largely of World Health Organization-recommended mass drug administration (MDA) of the anthelminthic drug praziquantel.^3^ MDA is recommended when the community prevalence of infection is above pre-set thresholds; however, there is a need for a more granular geographic understanding of disease burden to help guide MDA programs. The paper-based hematuria assays used to determine community prevalence are effective when results are aggregated, but they have insufficient sensitivity and specificity to diagnose individuals. Screening strategies with tools such as urine filtration and centrifugation for *S. haematobium* are time consuming, require skilled labor, and rely on functional laboratories that may be distant from the site of sample collection. Newer diagnostics include assays to detect circulating anodic antigen (CAA) and circulating cathodic antigen (CCA). However, CAA testing still requires considerable laboratory infrastructure,^4,5^ and CCA testing yields insufficient diagnostic performance for *S. haematobium*.^5,6^

Urine microscopy is the most widely available diagnostic technique with low enough type I and type II error to diagnose and screen for *S. haematobium*, but several factors limit its widespread use. First, because the eggs in urine samples may be in concentrations of 1 egg/mL or less,^7^ samples must undergo filtration,^8^ sedimentation,^9^ or centrifugation^10^ to increase their concentration prior to imaging on a conventional microscope. The sample preparation and imaging process is time-consuming and requires a trained technician. Second, the required resources for conventional microscopy are typically available in regional public health labs or hospitals in larger urban centers, making them less accessible to most people living in rural areas of endemic regions. Even in regions served by public health labs, the requisite time and transportation make conventional microscopy an impractical technique for individual diagnoses.

Here, we address these challenges by developing a mobile phone-based microscope (the “SchistoScope”) using compact, reversed lens optics.^11^ We modified a previous device, the LoaScope, to incorporate brightfield and darkfield imaging and to accept a specialized cartridge the captures and concentrates *S. haematobium* eggs for imaging by the SchistoScope. After validating the technology in laboratory testing, we evaluated the sensitivity and specificity of this handheld, point-of-care microscope in a pilot study in Ghana and Côte d’Ivoire.

## Results

### Isolation of *S. haematobium* eggs with a novel cartridge design

While mobile phone microscopy has sufficient resolution to resolve *S. haematobium* eggs (∼150 µm in length),^12^ a sample preparation protocol simple and efficient enough to be performed at the point of care has been lacking. Normally, sample preparation involves at least two steps: concentration of eggs into a small volume and transfer of that volume onto a glass slide or other sample holder for imaging. We aimed to develop a single-step sample preparation protocol that concentrated a significant portion of the eggs from a 10 mL urine sample into the ∼1 µL in-focus volume of the SchistoScope. We achieved this by producing a plastic cartridge that serves both as the filter and the slide for imaging. The diagnostic workflow using the cartridge and a mobile phone microscope is shown in Figure 1A-C.

**Figure 1:**
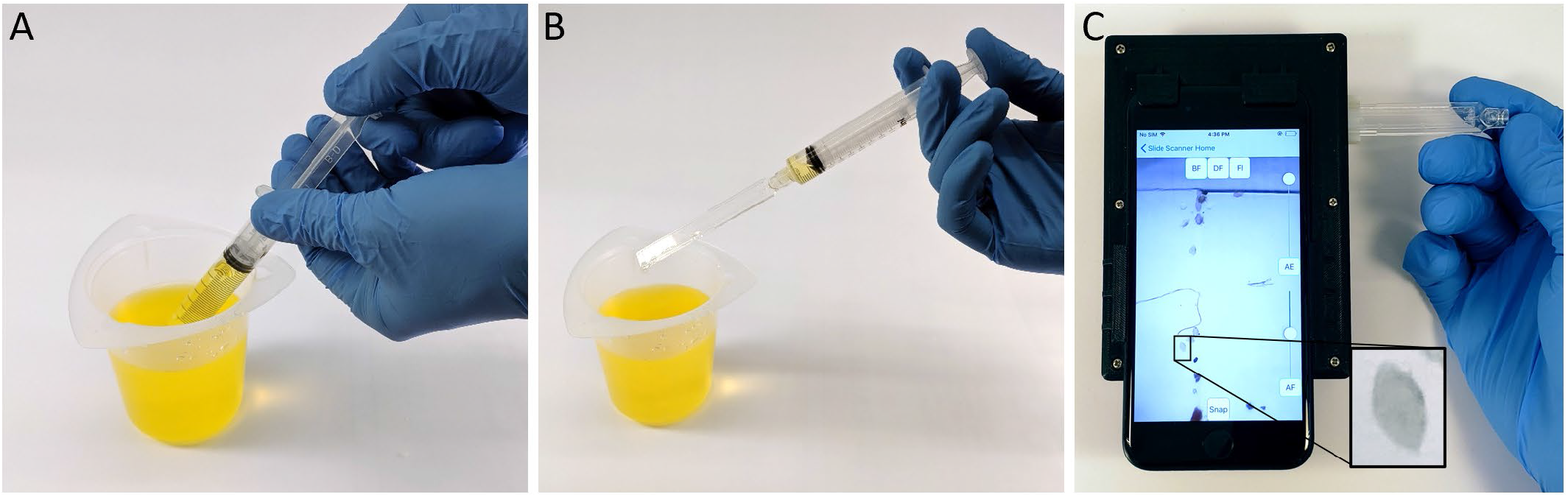
Diagnostic workflow for *S. haematobium* eggs using the SchistoScope. (A) Urine is drawn from a collection cup into a syringe. (B) The cartridge is attached to the syringe and the urine is pushed through, trapping the eggs. (C) The cartridge is inserted in the SchistoScope, where microscopic images of eggs are captured by the phone’s camera, coupled to an external reversed lens.

To concentrate *S. haematobium* eggs for imaging, the cartridge is designed with a port at one end to attach to a syringe containing 10 mL of urine and an outlet at the other end, such that the urine can be pushed through the single linear channel within the cartridge (Figure 2A). The channel is 3 mm wide and decreases from 200 µm to 20 µm tall over a 30 mm length, allowing particles of decreasing size to be captured against the clear, flat, top face for imaging (Figure 2B). Particles larger than 200 µm are prevented from entering the channel, while those below 20 µm pass through the outlet. The gradient in channel height helps to spatially separate *S. haematobium* eggs from other contaminants in urine.

**Figure 2:**
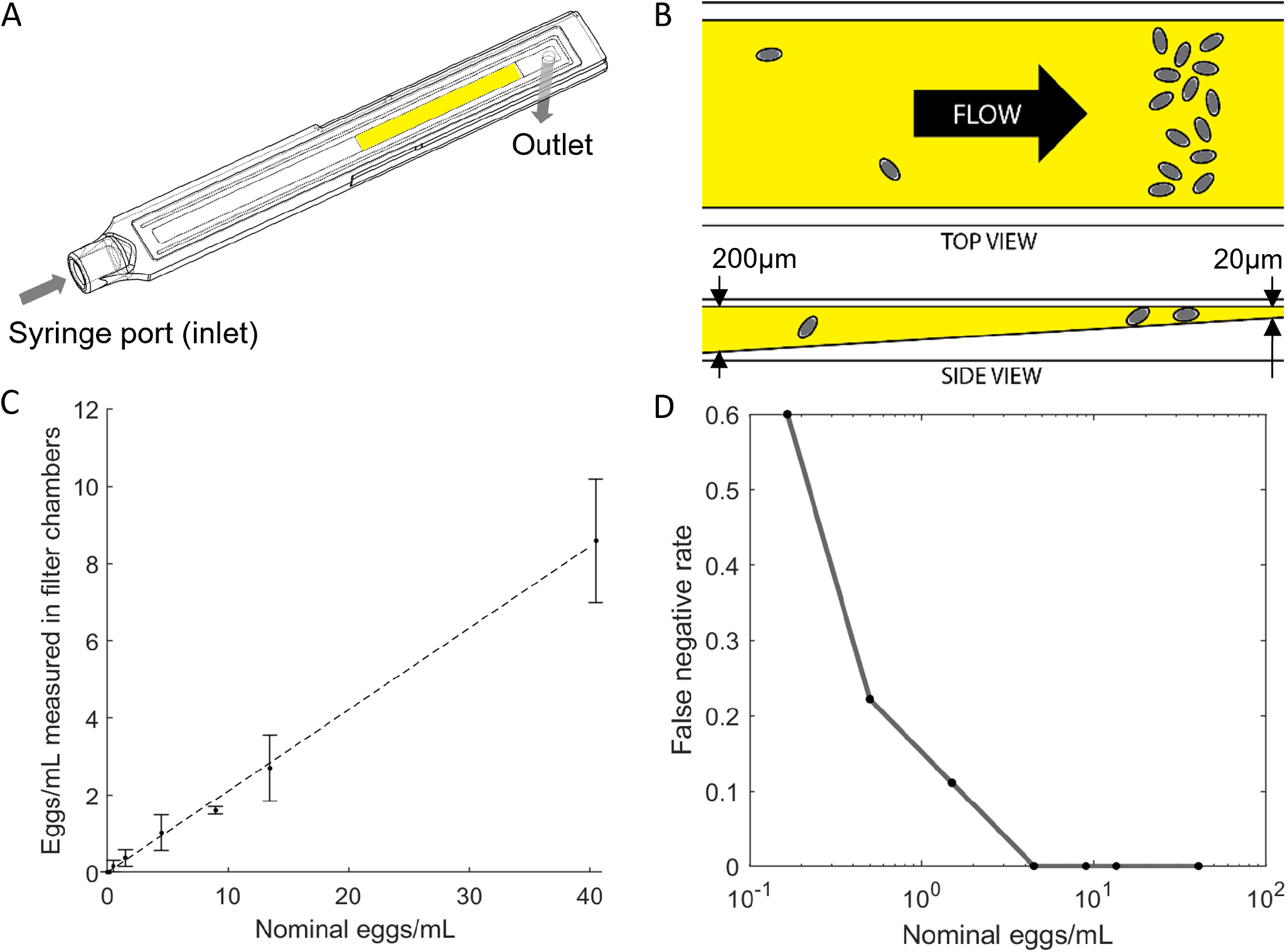
Design and laboratory testing of the *S. haematobium* egg capture cartridges. (A) Wireframe drawing of the cartridge. (B) Cartoon of the channel that captures eggs to be imaged inside the cartridge (not drawn to scale). (C) Manual counts of eggs captured in cartridges from titrated solutions of *S. haematobium* eggs in saline. The dotted line represents 21% of nominal egg concentrations. (D) Measured false negative rate of sample preparation process (no eggs captured).

In laboratory tests, the cartridges consistently capture around 21% of *S. haematobium* eggs resuspended in 10 mL of saline solution within the SchistoScope field of view (Figure 2C), based on eggs counted manually from SchistoScope images. In field experiments, this capture rate was comparable to the traditional filtration and imaging. The remaining 79% of eggs are either caught on the plastic surfaces of the collection cup, the syringe, or in a small trough that runs along the perimeter of the channel and protects the channel during solvent bonding of the cartridges. (A future iteration of the cartridge design will eliminate the trough.) Cartridges that contain no visible eggs from the prepared solutions of eggs in saline are considered a false negative produced by the sample preparation process. The measured rate of these occurrences is shown for varying egg concentrations in Figure 2D. By this metric, 100% sensitivity for the sample preparation process is observed at concentrations above 4.5 egg/mL and 89% sensitivity above 1.5 eggs/mL.

### Imaging of *S. haematobium* eggs with the SchistoScope

The SchistoScope was adapted from a previous mobile phone-based diagnostic device,^13,14^ where it was used to detect *Loa loa* microfilaria in peripheral whole blood. This device uses the built-in CMOS sensor and lens of a mobile phone, coupled to an additional lens on the outside of the phone. The additional lens is identical to the built-in lens, but reversed, creating a microscope where a pixel-sized area on the sample is imaged directly onto a pixel of the phone’s camera sensor. The reversed lens approach to mobile microscopy has the advantage of providing highly corrected optics over a large field-of-view and costs only a few US dollars to produce. The result of this configuration is a microscope with <5 µm resolution over the 12 mm^2^ area required to image the majority of the schistosome eggs captured in the cartridge.

The device was altered to use an Apple iPhone 8 for its smaller pixel pitch and faster processor compared to the iPhone 5S used in the original device. The microscope is easily adaptable to other phone models in future iterations.

### Pilot studies with the SchistoScope in Ghana and Côte d’Ivoire

The SchistoScope was evaluated in two pilot studies of school-age children in *S. haematobium* endemic areas. 63 patients from Sorodofo-Abaasa town, north of Cape Coast, Ghana and 142 patients from Azaguié region of southern Côte d’Ivoire provided urine samples as part of surveillance for an ongoing MDA program. Locations of the field studies are shown in Figure 3A. There was a combined 55% prevalence of *S. haematobium* in these urine samples. Additionally, a significant amount of debris was visualized, which was not present in the reconstituted samples used for bench testing. The debris included clothing fibers, mites, blood, crystals, and epithelial tissue (Figure 3B).

**Figure 3:**
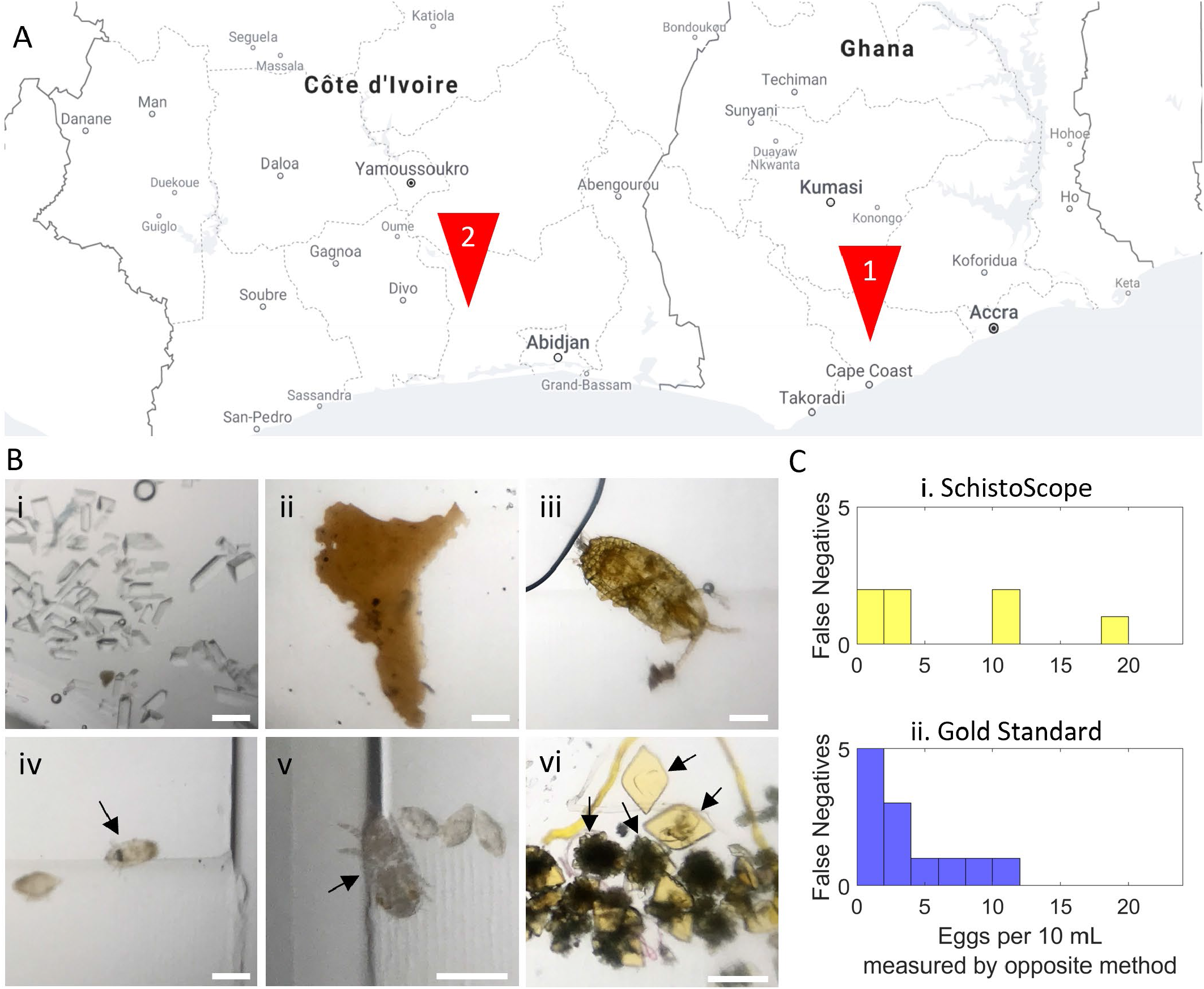
Field testing of the SchistoScope for *S. haematobium* egg detection. (A) Two sites were chosen for field studies: Sorodofo-Abaasa town, north of Cape Coast, Ghana, and the Azaguié region of southern Côte d’Ivoire. (B) Examples of additional objects captured by the cartridges and imaged by the SchistoScope. (i) Struvite crystals (ii) Epithelial tissue (iii) Unidentified mite (iv) Schistosome miracidium (v) Unidentified mite (vi) Uric acid crystals. All scale bars = 200µm (C) False negative rates of the diagnostic techniques for the population of 205 patients with 55% *S. haematobium* prevalence (i) The measured false negative rate of the SchistoScope using field gold standard technique as ground truth. (ii) The measured false negative rate of the field gold standard technique (filtration followed by conventional microscopy) using the SchistoScope as ground truth. Eggs were counted manually in both cases. False negatives were counted as instances where one method isolated at least one egg and the other did not.

The SchistoScope demonstrated a sensitivity of 90.9% and specificity of 91.1% for *S. haematobium* diagnoses compared to the field gold standard, consisting of conventional urine processing and light microscopy. The mismatch between the SchistoScope and gold standard is presented in Figure 3C, where false negatives of the two techniques are summed for different egg concentrations, using the opposite method as ground truth information for concentration. Overall, the SchistoScope produced comparable or slightly fewer false negatives compared to the gold standard.

### Automated detection of *S. haematobium* eggs using machine learning

We used machine learning algorithms for *S. haematobium* egg detection from images captured using the SchistoScope on samples collected in field settings. A number of different algorithms have been developed for object detection on a mobile device.^15,16^ As a starting point, we chose to compare a set of popular object detection architectures for egg detection. Using transfer learning we compared RetinaNet,^17^ MobileNet,^18^ and EfficientDet^19^ architectures that had been trained on the COCO 2017 dataset,^20^ retaining the feature extraction layers and fine tuning the dense layers of these models to detect *S. haematobium* eggs as a single class. We evaluated model performance at detecting eggs in the patient data (see Materials and Methods).

Detected regions within an image by each architecture are enclosed by bounding boxes and returned with a probability of containing the desired class, which we refer to as the detection score. Objects having a detection score above a certain threshold are deemed positive for the desired class. We evaluated the influence of the detection threshold on the sensitivity and specificity of the diagnostic for the RetinaNet architecture (Materials and Methods, Figure 4A). A detection threshold of 55% resulted in the best compromise between sensitivity (86%) and specificity (80%), and therefore we used this threshold to compare and contrast the performance of the RetinaNet architecture with different model dimensions, MobileNet and EfficientDet. We first tested the performance of RetinaNet implemented with different numbers of residual layers (ResNet-50, ResNet-101, ResNet-152^21^). The RetinaNet architecture had improved egg detection performance with a higher number of residual layers, with values coming close to those obtained by the trained user with manual egg counting (ResNet-101 sensitivity 91%, specificity 85%, ResNet-152 sensitivity 82%, specificity 90%, Figure 4B). In contrast, our implementation of the MobileNet and EfficientDet architectures were comparatively not as sensitive or specific (MobileNet sensitivity 77%, specificity 70%, EfficientDet sensitivity 32%, specificity 65%).

**Figure 4:**
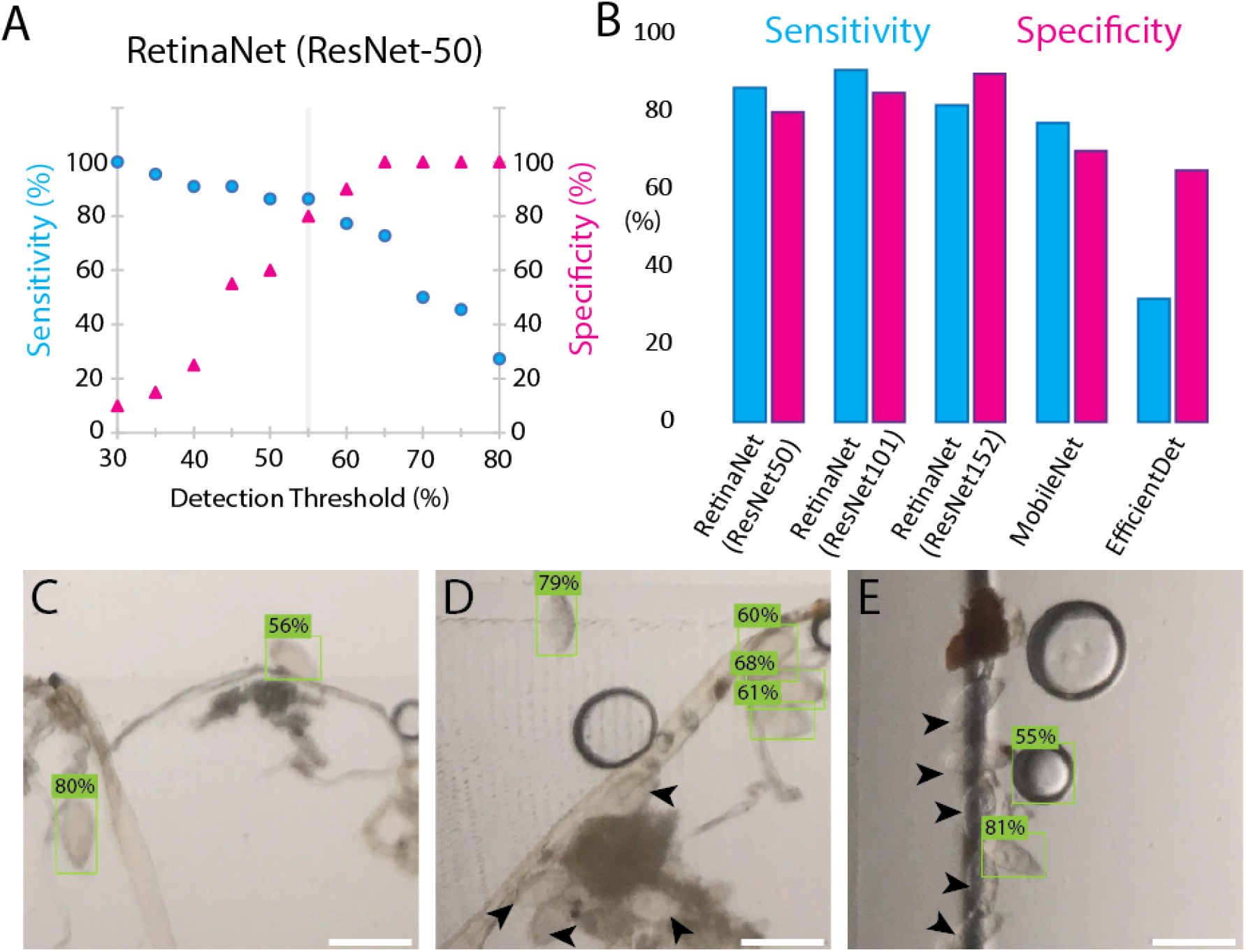
Automated *S. haematobium* egg detection using transfer learning. (A) By varying the detection threshold for a single region of interest, the sensitivity (blue dots) and specificity (magenta triangles) of the algorithm as a whole can be tuned. The example is shown for RetinaNet (ResNet-50) with the optimal detection threshold at 55%. (B) The ability of different neural networks to detect the presence of schistosome eggs in an image is quantified using sensitivity and specificity, with expert counts as the ground truth. (C-D) The algorithm performs well at detecting isolated eggs and rejecting other debris from a crowded field of view, but does not identify eggs that are clustered with debris (black arrowheads). (E) At the edges of the cartridge, many eggs are not identified by the algorithm. In this image, an air bubble is incorrectly classified as an egg, albeit with lower probability. All scale bars = 200 µm.

Although the machine learning algorithms could achieve reasonable sensitivity and specificity, the total eggs counted by all of the models tested was poor, capturing only 40-50% of the total eggs labelled in the ground truth. To investigate this further, we examined instances where the algorithm failed to identify eggs correctly within a sample. There were two main instances when eggs were not detected by the algorithm (Figure 4C-E). First, in very high load samples, eggs sometimes aggregated in large clumps with other debris, causing the algorithms to miss them. Second, in these high load samples, eggs could become stuck at the edge of the channel wall, also causing the algorithm to miss them. The algorithm performed best at detecting single isolated eggs, which made up most of the patient and training data. Even in the high load instances, the sample often (∼98% of cases) contained eggs that were well separated and positioned in the sample channel for detection. Therefore, although egg clumping and edge effects impacted the overall egg count accuracy, it had a limited impact on the overall sensitivity and specificity of the algorithms. This suggests that machine learning can be an effective method for analyzing SchistoScope images, especially as larger training datasets are collected.

## Discussion

The lack of high-performance, rapid, point-of-care diagnostic and screening tools for *S. haematobium* presents a barrier to deworming efforts in endemic settings. Diagnosis of individuals, rather than local populations, is especially vital for regions that have relatively mild worm burdens, often where mass drug administration (MDA) campaigns are active. In such settings, high-sensitivity diagnostics are imperative to implement individual test-and-treat control measures and are also useful to rapidly screen regions to determine if prevalence thresholds favor MDA. Additionally, wide-scale administration of praziquantel may contribute to the development of drug resistance, and more judicious use of MDA programs in carefully screened regions may minimize the chances of this developing.^22^

Here, we have presented inexpensive and effective techniques for sample preparation, imaging, and analysis of samples. To simplify the sample preparation protocol and enable *S. haematobium* egg concentration at the point of care, we developed a cartridge with the ability to collect and concentrate schistosome eggs from low concentration solutions in urine. After loading, the cartridge is then automatically imaged by the SchistoScope, followed by egg identification and quantification by expert technicians or a machine learning algorithm. With further improvements in cartridge manufacturing and an expanded training set for the machine learning algorithm, the SchistoScope could provide a rapid and effective strategy for individual-level quantitative diagnosis for *S. haematobium*. The high sensitivity and specificity yielded by the technique in these early field studies are promising for the viability of the technique to provide a new point-of-care test of *S. haematobium*, and to justify additional studies with optimization and thousands of patient samples.

Using the cartridge to concentrate *S. haematobium* eggs for microscopic imaging significantly reduces the complexity of sample preparation compared to existing field filtration and centrifugation techniques and likely represents a very time efficient manner to process samples at the point of sample collection. Similarly, a mobile phone-based microscope offers a cost-effective method to conduct surveillance that can be performed without the need for a regional laboratory.^23^ Like the existing point-of-care diagnostics, the consumable products (cup, syringe, cartridge) are sterile and disposable. The cost of materials at volume is expected to be less than 1 USD, which compares well with existing diagnostic techniques.

Multiple diagnostic tests have been used to detect schistosomiasis, with a range of performance, cost, and availability (Table 1). Compared to conventional light microscopy as the field gold standard, the SchistoScope demonstrated a sensitivity of 90.9% and specificity of 91.1% for the manual detection of *S. haematobium* eggs. It is important to note, however, that gold standard measurements do not have perfect specificity, especially when conducted on a single sample by a single microscopist. Most (or all) of the SchistoScope samples that classified as “false positives” (i.e. determined by an expert to be positive on the SchistoScope but found to be negative using the field gold standard) were likely true positives that were simply missed by the gold standard preparation.^24^ Cartridges with these samples were subsequently analyzed under light microscopy to validate the true presence of *S. haematobium* eggs, so the true specificity of the SchistoScope is likely comparable to, or better than, the traditional sample preparation and evaluation via light microscopy.

**Table 1:**
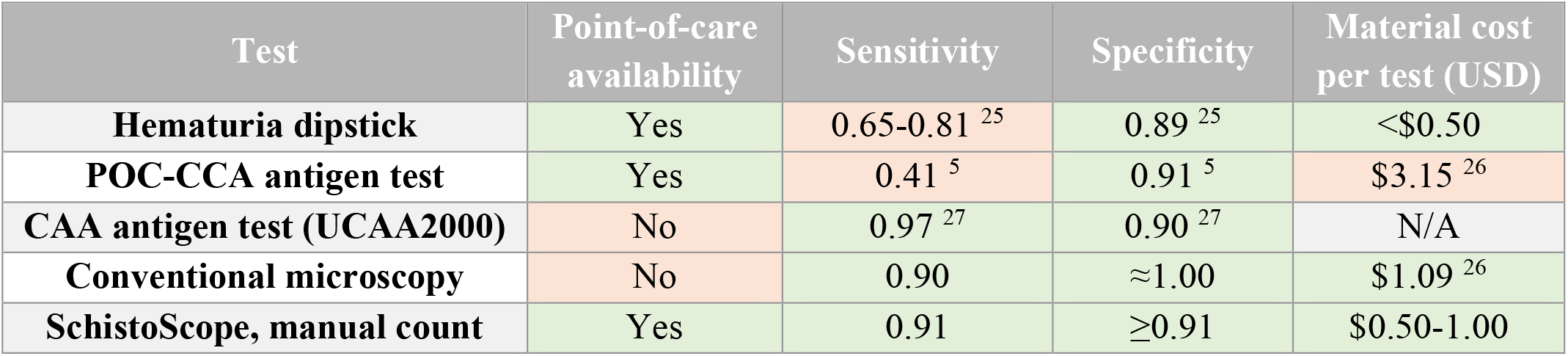
Comparison of existing diagnostics for S. haematobium, including the mobile phone microscopy technique reported here. The CAA antigen test is not commercially available at the time of this writing.

One strategy to improve the efficiency of screening at the point of sample collection is to automate the analysis of images on the mobile phone device, especially considering the paucity of trained laboratory technicians and microbiologists in rural areas where schistosomiasis is endemic. Machine learning algorithms for object detection provide a precise way to detect and classify complex objects in images, which can remove the need for an expert user for data analysis at the point of care. Training a machine learning algorithm from scratch requires a large amount of training data labelled by an expert in order to obtain a high accuracy model. To explore the possibility of using machine learning to detect *S. haematobium* eggs, we used transfer learning of pre-existing object detection models, retaining their feature detection layers and fine tuning the dense layers for egg detection. This strategy requires significantly less training data and could therefore be implemented using our current dataset (∼200 patient samples). The sensitivity and specificity that we obtained with this approach were close to, albeit less than, the values obtained by a trained expert with manual egg detection. Our implementation of the RetinaNet (ResNet 101) architecture yielded the best results with a specificity of 91% and sensitivity of 85%. This network also ran rapidly on a Pixel4 mobile device taking ∼6 seconds to perform inference on a 640×640 pixel region. The total time to perform the diagnostic test is therefore reasonable for a point of care diagnostic, returning a positive or negative result within minutes.

Taken together, the high sensitivity and specificity of the machine learning algorithm combined with the fast inference time show that a machine learning algorithm can substitute for a trained technician at the point of contact, should a technician not be available. With the addition of more training data obtained in future field studies, we anticipate that a fully trained model would have increased specificity and sensitivity versus the current model implemented using transfer learning, with feature extraction layers that are specific for the egg detection problem. In addition, the current algorithm has been optimized for the detection of a single class (*S. haematobium* eggs), but could easily be retrained to detect other classes such as epithelial tissue or various types of crystals that were present in the field-collected samples and are either debris or hallmarks of other pathologies (Figure 3B). A multiclass object detection model of this type has the potential to diagnose different diseases based on our simple sample preparation and imaging platform. Furthermore, the use of multiple contrast methods, such as dark field and brightfield, to detect S. haematobium eggs has the potential to improve object detection accuracy.

Looking ahead, the general strategy of inexpensive size-based filtration coupled with mobile phone imaging is promising. The addition of flow and filtration steps in the protocol described here may open the door to point-of-care diagnostics for additional diseases. An obvious next target is *Schistosoma mansoni* and soil-transmitted helminths, which share large regions of coendemicity with *S. haematobium*. Conveniently, *S. mansoni* is treated with the same drug, praziquantel.^8^ Because the eggs of *S. mansoni* are primarily shed through stool rather than urine, formation of a fecal slurry and upstream filtration before capturing eggs in the cartridges would likely be necessary. Combined with further advances in machine learning algorithms, the use of a compact mobile phone-based microscope with disease-specific cartridges has the potential to address multiple disease diagnostic needs with a single device.

## Materials and Methods

### Cartridge

The cartridges were produced in two halves: one half including the bottom and side faces of the channel, linked to a syringe port and an outlet hole. Around the channel is a flat datum surface to locate the second half of the channel during bonding. Within the datum surface, there is a small recess that runs along the perimeter of the channel, to prevent the propagation of solvent during bonding. This half of the cartridge was injection molded in clear polycarbonate. The second half of the cartridge is a flat piece laser cut from 600 µm thick clear polymethylmethacrylate. Solvent bonding was achieved by applying a drop of dichloromethane at the seam between the two parts while the parts are held in contact. After the solvent propagates into the seam, the parts are held in contact for 30 seconds and left in ambient conditions for 48 hours. After 48 hours, the parts were flushed with deionized water from a syringe and dried with a stream of air. Bench testing of the cartridges was performed using *S. haematobium* eggs provided by the NIH-NIAID Schistosomiasis Resource Center, which were isolated with hamsters. The eggs were diluted into 1x PBS and mixed by inversion before 10 mL were pushed through each test cartridge.

### Machine learning

We split the field data into a training and test set to train and evaluate the performance of different object detection algorithms. Initial image pre-processing was added to the analysis pipeline and consisted of the following steps. The field of view of the phone covers the entire width of the cartridge channel. Images were cropped to regions of interest of 640×640 pixels and converted to RGB jpeg format. In some instances, there were differences in the white balance of the images collected from different devices which were normalized for throughout the training and test sets by scaling the intensity of the green color channel by 5-8%. The training set was then labelled for instances of eggs with bounding boxes and used to for transfer learning. Transfer learning was implemented using TensorFlow 2 object detection API and Keras using models trained on the COCO 2017 dataset.^16,20^ The feature extraction layers of the different models were retained and the weights of the dense layers fine-tuned for egg detection. Object detection models such as RetinaNet have separate networks for classification and bounding box regression. In our implementation, we only retrained the dense layers of the classification network for a single class. As part of the model training, we added data augmentation including random flipping and rotation of the images, and random hue, contrast, saturation and brightness adjustments. Training was implemented with a batch size equal to the number of training images and a learning rate of 0.01 using the stochastic gradient descent optimizer with momentum set to 0.9. To evaluate the performance of the trained network on the mobile device we converted the models to the TensorFlow Lite format and implemented inference on the phone using Android Studio. Values for the inference speed were evaluated on a 640×640 frame being streamed from the mobile phone camera.

We used three different metrics to evaluate the performance of the different model architectures. True positives, true negatives, false positives and true positives were evaluated at the image level. The values for sensitivity and specificity therefore represent that of the diagnostic. In addition, we quantified the total percentage of eggs that was correctly detected by each of the models across the entire test set

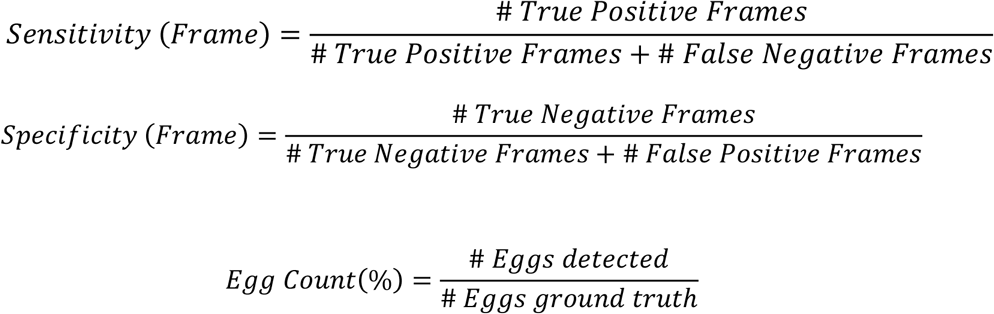

### Sample collection and field protocol

This study was integrated with pre-existing studies and control efforts, and institutional review board approval was granted; CNESVS #IRB000111917 (Côte d’Ivoire), UCCIRB/EXT/2017/33 (University of Cape Cost, Ghana), REB 14-8128 (University Health Network, Toronto, Canada). Urine was collected between 10:00 and 14:00 and processed the same day. Urine samples were first shaken, and then 20 cc was removed; 10 cc for evaluation by conventional microscopy and 10 cc by the SchistoScope. For conventional microscopy, 10 cc was pressed via a syringe through filter with 20 μm pores, and the filter was then removed, placed on a glass slide with a drop of Lugol’s iodine, and evaluated under 20x and 40x lenses by a trained laboratory technician. 10% of samples were randomly selected for quality control by a microbiologist.

The other 10 cc of urine was pressed via a syringe through the cartridge over 10 seconds, with care to eliminate air bubbles in the syringe. 10 seconds was chosen to avoid extreme pressure on the syringe; increasing the flow rate dramatically can elastically deform the plastic in the cartridge window, while increasing the drag force on the eggs, allowing eggs to escape through the outlet. The total time for the sample preparation, from a cup of urine, to a microscopy-ready cartridge was about 30 seconds per sample.

### Statistical analysis of field data

In the field study, we estimated the sensitivity and specificity of the SchistoScope, with visual interpretation, compared with conventional light microscopy serving as a reference standard. We calculated exact binomial 95% confidence intervals for each metric. All analyses were performed using R (version 4.0.5).

## Data Availability

Data used in this study is available from the corresponding author upon reasonable request.

## Acknowledgements

We thank members of the Fletcher Lab for helpful discussions. We appreciate the samples generously provided by the NIH-NIAID Schistosomiasis Resource Center. We thank the participants in Sorodofo-Abaasa town, Ghana, and the Azaguié region of Côte d’Ivoire.

